# SUCCESSFUL MANUFACTURING OF CLINICAL-GRADE SARS-CoV-2 SPECIFIC T CELLS FOR ADOPTIVE CELL THERAPY

**DOI:** 10.1101/2020.04.24.20077487

**Authors:** Wing Leung, Teck Guan Soh, Yeh Ching Linn, Jenny Guek-Hong Low, Jiashen Loh, Marieta Chan, Wee Joo Chng, Liang Piu Koh, Michelle Li-Mei Poon, King Pan Ng, Chik Hong Kuick, Thuan Tong Tan, Lip Kun Tan, Michaela Su-fern Seng

**Affiliations:** Department of Haematology/Oncology, KK Hospital, SingHealth, Singapore; Duke-NUS Medical School, Singapore; Department of Haematology-Oncology, National University Hospital, Singapore; Department of Haematology, Singapore General Hospital, Singapore; Department of Infectious Disease, Singapore General Hospital, Singapore; Department of Infectious Disease, Sengkang General Hospital, Singapore; Blood Services Group, Health Sciences Authority, Singapore; Yong Loo Lin School of Medicine, National University of Singapore, Singapore; Department of Pathology and Laboratory Medicine, KK Hospital, SingHealth, Singapore

## Abstract

**Background:** Adoptive therapy with SARS-CoV-2 specific T cells for COVID-19 has not been reported. The feasibility of rapid clinical-grade manufacturing of virus-specific T cells from convalescent donors has not been demonstrated for this or prior pandemics.

**Methods:** One unit of whole blood was collected from each convalescent donor following standard blood bank practices. After the plasma was separated and stored separately, the leukocytes were stimulated using overlapping peptides of SARS-CoV-2, covering the immunodominant sequence domains of the S protein and the complete sequence of the N and M proteins. Thereaftesr, functionally reactive cells were enriched overnight using an automated device capturing IFNγ-secreting cells.

**Findings:** From 1×10^9^ leukocytes, 0.56 to 1.16×10^6^ IFNγ+ T cells were produced from each of the first two donors. Most of the T cells (64% to 71%) were IFNγ+, with preferential enrichment of CD56+ T cells, effector memory T cells, and effector memory RA+ T cells. TCRVβ spectratyping revealed oligoclonal distribution, with over-representation of subfamilies including Vβ3, Vβ16 and Vβ17. With just two donors, the probability that a recipient in the same ethnic group would share at least one donor HLA allele or one haplotype could be as high as >90% and >30%, respectively.

**Interpretations:** This study is limited by small number of donors and absence of recipient data; however, crucial first proof-of-principle data are provided demonstrating the feasibility of clinical-grade production of SARS-CoV-2 specific T cells for urgent clinical use, conceivably with plasma therapy concurrently. Our data showing that virus-specific T cells can be detected easily after brief stimulation with SARS-CoV-2 specific peptides suggest that a parallel diagnostic assay can be developed alongside serology testing.

**Funding:** The study was funded by a SingHealth Duke-NUS Academic Medicine COVID-19 Rapid Response Research Grant.

## INTRODUCTION

A novel severe acute respiratory syndrome coronavirus 2 (SARS-CoV-2) is the cause of Coronavirus Disease 2019 (COVID-19).^1-2^ No specific treatment has been proven to be effective. While vaccines are being developed, passive immunity can be acquired immediately by infusion of plasma from convalescent donors into newly infected patients.^3^ Currently after plasma is extracted from the donor’s blood unit, the white blood cells are discarded. We hypothesize that these white blood cells contain SARS-CoV-2 specific T cells which can be used for adoptive therapy.

The aim of this study is to demonstrate the feasibility of preparing clinical-grade SARS-CoV-2 specific T cells from convalescent donors for urgent clinical use. The specific hypothesis is that SARS-CoV-2 specific T cells can be isolated from the blood of convalescent donors rapidly and efficiently using SARS-CoV-2 specific peptides and an automated medical device for emergent treatment of severe COVID-19 disease.

## METHODS

### Donors

Convalescent donors were referred by their infectious disease physicians. Eligibility criteria included age 21 to 65, a history of COVID-19 with documented positive test for SARS-CoV-2 in the past but the test had become negative since, fulfillment of all blood donation criteria per standard blood bank practices, and an informed consent with approval by the hospital Institutional Review Board (NCT04351659). One unit of whole blood was collected from each donor once. The medical history of the first two donors is summarized in **Supplemental Table 1**.

**Table 1.** Donor Lymphocyte Compositions Before and After Enrichment

|  | Donor 1 |  |  | Donor 2 |  |  |
| --- | --- | --- | --- | --- | --- | --- |
|  | Pre-Selection | Negative<br>Fraction | Positive<br>Fraction | Pre-Selection | Negative<br>Fraction | Positive<br>Fraction |
| Lymphocyte subsets |  |  |  |  |  |  |
| T cells | 86% | 71% | 71% | 72% | 73% | 58% |
| B cells | 7% | 11% | 25% | 22% | 22% | 38% |
| NK cells | 7% | 18% | 4% | 6% | 5% | 4% |
| T cells |  |  |  |  |  |  |
| CD4:CD8 | 65:35 | 61:39 | 75:25 | 62:38 | 61:39 | 77:23 |
| CD56<sup>-</sup>:CD56<sup>+</sup> | 97:3 | 96:4 | 78:22 | 94:4 | 92:8 | 83:17 |
| Naïve | 23.2% | 17.7% | 8.5% | 33.4% | 33.0% | 15.9% |
| CM | 34.9% | 35.6% | 19.5% | 40.8% | 42.5% | 23.0% |
| EM | 35.7% | 39.2% | **67.4%** | 18.7% | 17.4% | **51.7%** |
| T<sub>EMRA</sub> | 6.3% | 7.5% | 4.6% | 7.1% | 7.1% | 9.4% |
| CD3<sup>+</sup> IFNγ<sup>+</sup> cells | | | | | | |
| Naïve | 1.9% | 1.8% | 0.2% | 12.7% | 13.2% | 1.5% |
| CM | 59.5% | 59.1% | 12.8% | 71.7% | 72.3% | 16.5% |
| EM | 37.9% | 38.0% | **81.9%** | 14.4% | 13.3% | **69.8%** |
| T<sub>EMRA</sub> | 0.8% | 0.3% | **5.0%** | 1.2% | 1.2% | **12.2%** |
| CD4<sup>+</sup> IFNγ<sup>+</sup> cells | | | | | | |
| Naïve | 1.6% | 1.9% | 0.3% | 10.8% | 11.9% | 1.5% |
| CM | 65.5% | 62.9% | 10.3% | 68.7% | 68.6% | 15.6% |
| EM | 32.6% | 35.0% | **89.4%** | 20.2% | 19.1% | **81.6%** |
| T<sub>EMRA</sub> | 0.3% | 0.2% | 0.0% | 0.3% | 0.4% | 1.4% |
| CD8<sup>+</sup> IFNγ<sup>+</sup> cells | | | | | | |
| Naïve | 6.4% | 0.0% | 0.2% | 24.0% | 24.6% | 0.8% |
| CM | 23.0% | 33.3% | 2.3% | 57.7% | 57.7% | 10.6% |
| EM | 60.3% | 66.7% | **78.7%** | 15.8% | 15.1% | **32.5%** |
| T<sub>EMRA</sub> | 10.3% | 0.0% | **18.8%** | 2.5% | 2.6% | **56.1%** |

### Cell processing

After plasma extraction and buffy coat preparation, the white cells were processed using the automated CliniMACS Prodigy IFN-γ Cytokine Capture System^®^ (CCS) (Miltenyi Biotec, Germany). Briefly, 1×10^9^ cells were stimulated using overlapping peptides of SARS-CoV-2, covering the immunodominant sequence domains of the S protein (GenBank MN908947.3, Protein QHD43416.1), and the complete sequence of the N and M proteins (GenBank MN908947.3, Protein QHD43423.2; GenBank MN908947.3, Protein QHD43419.1).^4^ The peptide pools were short 15-mer peptides with 11-amino-acid overlaps, which can bind to MHC class I and class II complexes and thus were able to stimulate both CD4+ and CD8+ T cells. Thereafter, the cells were labeled with the Catchmatrix Reagent containing bispecific antibodies for CD45 and IFN-γ, which was secreted by the stimulated target cells during the secretion period. After the secretion phase, the cell surface-bound IFN-γ was targeted by the Enrichment Reagent, which contained IFN-γ-specific antibody conjugated to superparamagnetic iron dextran particles (MACS^®^ MicroBeads), thus allowing subsequent immunomagnetic cell separation. The unlabeled cells (CCS negative fraction) passed through the built-in magnetic column, whereas the Microbead-labeled cells (CCS positive fraction) were retained in the magnetic field. Afterwards, the magnetic field was turned off and the target cells were eluted into the target cell bag. The entire process took only 12 hours for cell manufacturing.

### Flow cytometry

For the analysis of cell compositions, two antibody panels were used. The first panel contained CD45-VioBlue, CD4-VioGreen, CD3-FITC, Anti-IFN-γ-PE, CD45RO-PEVio770, CD62L-APC, CD8 APCVio770, and 7-AAD for product release and assessment of cell subsets that were IFN-γ positive. The second panel contained CD45-VioBlue, CD4-VioGreen, CD3-FITC, CD16-PE, CD56-PE, CD19-PEVio770, CD14-APC, and CD8-APCVio770 for the assessment of cell content including B cells, NK cells, CD56+ T cells and CD4:CD8 ratio. Cell acquisition and analyses were performed using the MACSQuant^®^ Analyzer 10 in combination with Express Modes CCS_Purity_h_01 and CCS_Immune_Cell_Composition_h_01 (Miltenyi Biotec, Germany).

### TCR Vβ spectratyping

The CDR3-encoding region of the TCRVβ gene was amplified using 24 TCRVβ subfamily-specific primers and a carboxyfluorescein (FAM)-conjugated TCRVβ constant region-specific primer.^5^ The PCR products were denatured with Hi-Di formamide (Applied Biosystems, Carlsbad, CA) and electrophoresed along with Gene Scan-600 LIZ size standard (Applied Biosystems) on a SeqStudio Genetic Analyzer (Applied Biosystems). The overall complexity of TCRVβ in each subset was calculated by summation of the total number of peaks in each subfamily.

## RESULTS

At the time of blood donation, the leukocyte count of Donor 1 was 7.33 and the lymphocyte count was 2.08 (×10^6^/ml). Of these, 1 ×10^9^ leukocytes were used for SARS-CoV-2 specific T cell manufacturing, yielding 1.16×10^6^ IFNγ+ T cells in the final product (positive fraction). Most of the CD3+ cells (74%) in the positive fraction were IFNγ+, in contrast to the low frequencies observed in the corresponding negative fractions (**Figure 1A**).

**Figure 1.**
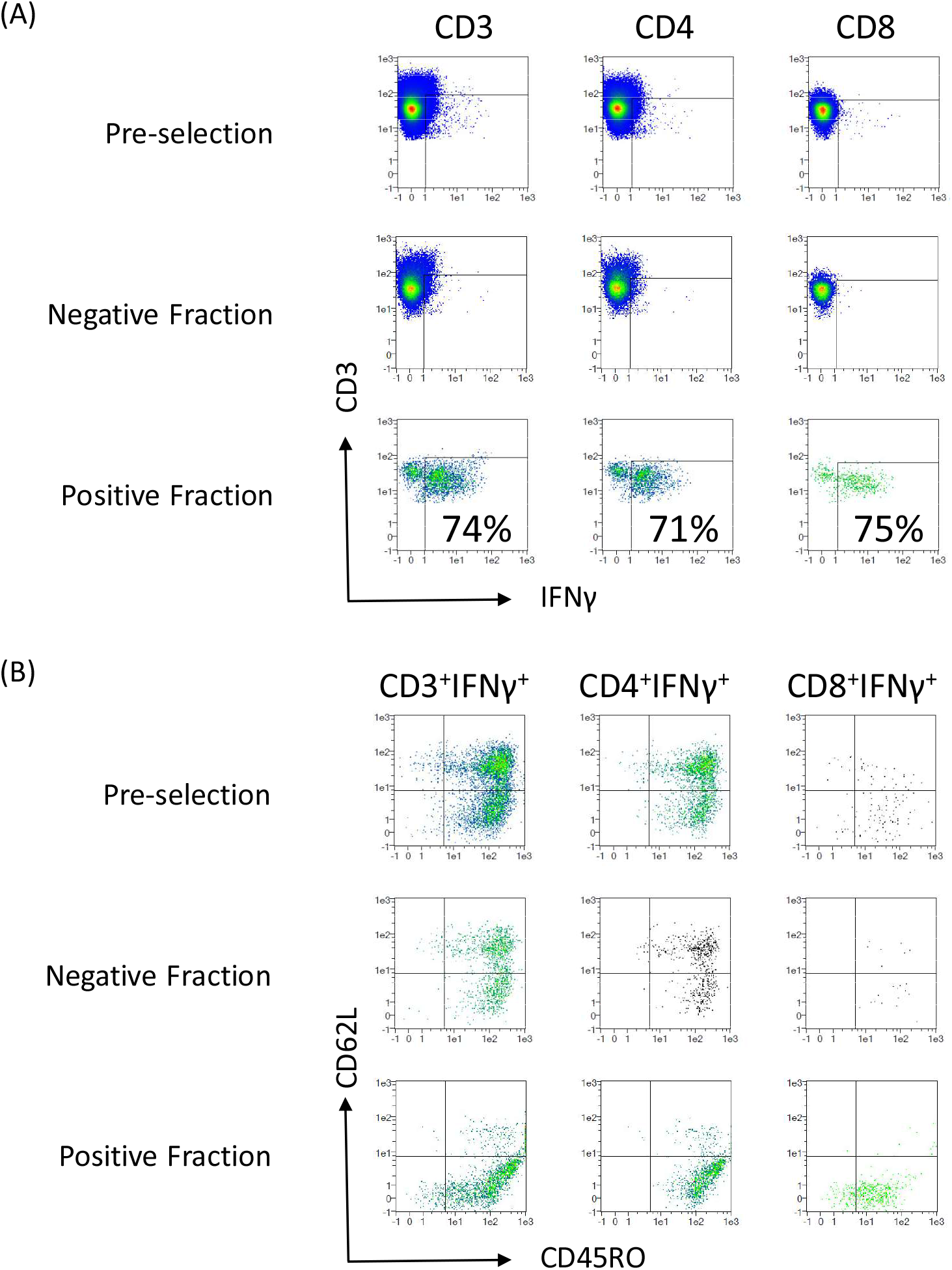
T cell immunophenotype before and after CCS enrichment. T cells from Donor 1 were assessed for IFNγ (A) and memory cell phenotypes (B) by flow cytometry before cell selection, post-enrichment in the positive fraction, and in the negative fraction. Separate analyses were performed for CD3+ cells, CD4+ cells and CD8+ cells. Panel (B) was gated on the IFNγ+ subsets showed in Panel (A).

### Changes in T-cell compositions before and after enrichment

Before CCS enrichment, the CD4:CD8 ratio was 65:35 and only 3% of the T cells were CD56+ (**Table 1**). The T naïve (TN), central memory (TCM), effector memory (TEM) and effector memory RA+ (TEMRA) cell ratio was 23:34:36:6. After CCS enrichment, the CD4:CD8 ratio increased to 75:25 and a substantial proportion of CD56+ T cells (22%) were observed. The TN, TCM, TEM and TEMRA ratio changed to 9:20:67:5, representing a relative depletion of naïve T cells and enrichment of effector memory cells. Gated on the IFNγ+ subpopulations, most (89%) of the CD4+ were TEM, while most of the CD8+ cells were TEM (79%) or TEMRA (19%). As expected, only 0.2% of the IFNγ+ T cells were naïve cells (**Figure 1B, Table 1**).

### TCRVβ usage

Before CCS isolation, polyclonal usage of TCRVβ in general Gaussian distribution was observed (**Figure 2**). After CCS enrichment, oligoclonal distribution was seen, and the Vβ score decreased from 142 to 43, with notable over-representation of Vβ3, Vβ16 and Vβ17. As expected, the TCRVβ usage in the negative fraction was similar to that before CCS isolation.

**Figure 2.**
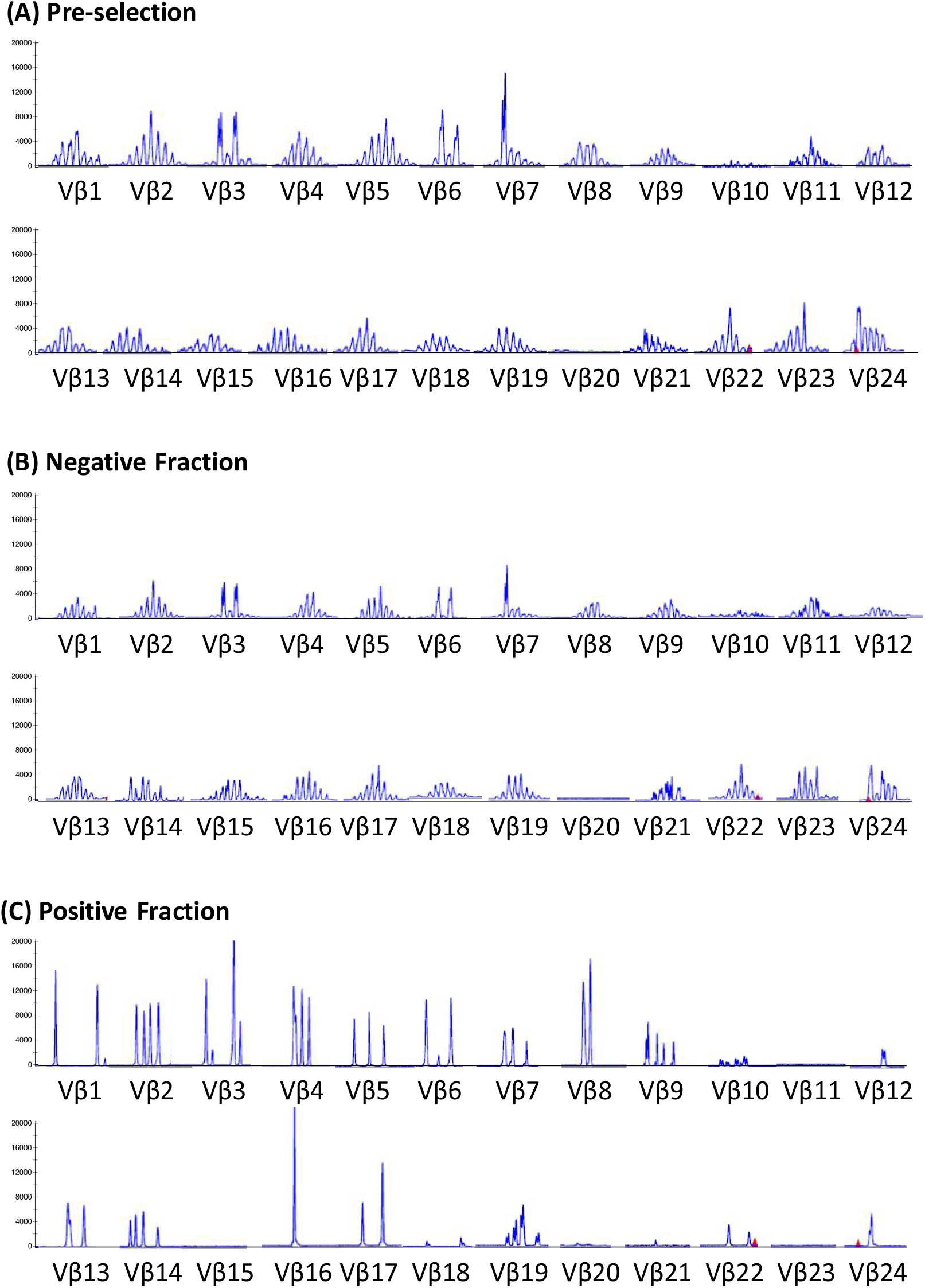
TCRVβ spectratyping. TCRVβ spectratyping for cells before enrichment, after enrichment in the positive fraction, and in the negative fraction.

### Reproducibility

To assess the reproducibility of CCS in preparation of SARS-CoV-2 specific T cells, another donor was recruited. In comparison with the first donor, the second donor had milder symptoms without lymphopenia during COVID-19 (**Supplemental Table 1**). After CCS processing identical to that of Donor 1, 64% of the T cells in the positive fraction were IFNγ+. In total, the positive fraction contained 0.56×10^6^ IFNγ+ T cells for clinical use.

Similar to Donor 1, the positive fraction from Donor 2 also had an increased CD4:CD8 ratio, relative enrichment of CD56+ T cells and restricted TCRVβ usage with a score 76. Likewise, most of the IFNγ+ cells were TEM in the CD4+ subset and TEM and TEMRA in the CD8+ population (**Table 1**).

### HLA repertoire

Both donors are Chinese Singapore residents and at least one of their HLA alleles is shared with >90% of the general Chinese population (**Supplemental Table 2**). The probability of sharing at least 3 alleles due to haplotype-sharing is >30%.

## DISCUSSION

COVID-19 is a rapidly progressing pandemic with no specific treatments or vaccines available thus far. We demonstrated herein for the first time that clinical-grade SARS-CoV-2 specific T cells can be isolated from the blood of convalescent donors rapidly and efficiently using SARS-CoV-2 specific peptides and an automated medical device for emergent treatment of severe COVID-19 disease. The manufacturing process is versatile and can produce cells overnight; thus, allowing flexible scheduling to avoid either shortage or wastage of volunteer blood cells. The availability of convalescent donors is naturally proportional to the size of pandemic.

In the majority of hospitalized patients with COVID-19, severe lymphopenia developed (lymphocyte count <0.8×10^9^/L), which correlated with disease severity;^6-9^ thus, candidates for adoptive T cell therapy may not require lymphodepleting preparative chemotherapy. Nevertheless, bidirectional alloreactivity is a concern. For graft-versus-host disease (GVHD), the CCS enriched product is depleted of naïve T cells, thereby limiting the risk of GVHD. In previous studies of HLA-mismatched virus-specific T-cell therapy, GVHD was not a barrier,^10-12^ as the clinically effective dose was as little as 5×10^3^ virus-specific T cells/kg, which was considerably lower than the clinical threshold for GVHD. For the T cells to be effective for virus control, they must act fast before complete donor-cell rejection. In this regard, the CCS system selects functionally rather than phenotypically and preferentially enriches those T cells that can secrete IFNγ within hours after stimulation with SARS-CoV-2 specific peptides by the nature of memory recall response. In contrast, de novo alloreactivity takes several days to develop, and could be longer in severely lymphopenic patients with severe COVID-19. Phenotypically, the CCS isolated cells are enriched for CD56+ T cells, CD4+ TEM cells, and CD8+ TEM and TEMRA cells, all of which are known to have potent and rapid pathogen response (thus captured preferentially by the CCS system).^5,13,14^

For pandemics caused by novel pathogens, development of specific vaccines for active immunization takes time; however, passive immunity can be acquired immediately via blood component transfusion from recovered patients. In this regard, promising preliminary data have been reported on the use of plasma therapy for COVID-19.^3^ Conceivably, our adoptive T-cell therapy approach can be easily combined with plasma infusion from the same donor or different donor. T cells are a “living drug” and can expand in vivo, which is in contrast pharmacokinetically to antibody, of which the level universally drops after infusion because of consumption or natural metabolism. Accordingly, a unit of donor’s blood may potentially treat more patients and the therapeutic effects of T cells may be more durable than that of plasma therapy. For a dose of 5×10^3^ SARS-CoV-2 T cells/kg, the cells from the donor with mild symptoms could provide sufficient cells for 3 adult recipients; correspondingly, the donor with severe symptoms could potentially benefit as many as 6 recipients. Pharmacodynamically, SARS-CoV-2 specific T cells may be used alone or synergistically with plasma therapy to establish immediately an adaptive immune status mimicking that after successful vaccination. Under the protection of adaptive immunity, the body will not depend solely on innate immune response, which may contribute to the cytokine storm and SARS pathogenesis.^15-17^ Coronavirus-specific T cells have been shown to be crucial for virus clearance and may dampen further damage to the lung by the dysregulated, overactive innate immunity.^17-20^

For the SARS-CoV-2 specific T cells to be effective, the donor cells must share some HLA with the recipient for proper viral antigen presentation to the donor TCRs. Previous studies have demonstrated that viral-specific T cells from donors who shared only one HLA were efficacious.^21^ With just two random donors, we estimated that more than 90% of the Singapore Chinese population will share at least one HLA allele with the manufactured cells (**Supplemental Table 2**),^22,23^ suggesting that our approach is feasible for both small-scale and large-scale, off-the-shelf delivery model and manufacture-on-demand model with simple ethnic-group matching.^24^ In 2018, Chinese made up 74.3% of the Singapore population. Our future goal will include recruiting some Malay and Indian donors into the pool as their ethnic groups constitute the other 13.4% and 9.0% of the Singapore population, respectively.^25^

This study is limited by small number of donors and absence of recipient data; therefore, product variability and clinical utility of SARS-CoV-2 specific T cells have yet to be determined. Nevertheless, we provide fundamental first proof-of-principle data herein demonstrating that it is feasible to produce clinical-grade SARS-CoV-2 specific T cells for urgent clinical use. These virus-specific T cells can be isolated from the blood of convalescent donors rapidly and efficiently using SARS-CoV-2 specific peptides and an automated medical device which is commonly available worldwide. The entire tubing-set is a closed system and thus reduces the requirements for cleanroom in lower-income regions. Extending from therapeutics to diagnostics, our data showing that virus-specific T cells can be detected easily after brief stimulation with SARS-CoV-2 specific peptides suggest that a diagnostic assay can be developed, particularly for patients similar to Donor 2 who had mild symptoms and might gone undiagnosed without a history of family contact.

## RESEARCH IN CONTEXT

### Evidence before this study

We searched PubMed on April 22, 2020, for articles that describe T-cell therapy for COVID-19, using the search terms “COVID-19”, “SARS-CoV-2”, or “2019-nCoV”, and “virus specific T cell therapy” or “adoptive T cell therapy”. No published work could be identified.

### Added value of this study

We demonstrated for the first time that clinical-grade SARS-CoV-2 specific T cells can be isolated from the blood of convalescent donors rapidly and efficiently using SARS-CoV-2 specific peptides and an automated medical device expedient for emergent treatment of severe COVID-19 disease. The cells were selected functionally rather than phenotypically and preferentially enriches those memory T cells that can secrete IFNγ within hours after stimulation with SARS-CoV-2 overlapping peptides.

### Implications of all the available evidence

By documenting that rapid production of clinical-grade SARS-CoV-2 specific T cells is feasible, our data encourage further study of these T cells for emergent treatment of COVID-19 disease until effective vaccines or treatment are developed. Our novel approach also sets a new paradigm for management and diagnosis in future pandemics with other novel pathogens.

## Data Availability

The corresponding authors had full access to all the data in the study and had final responsibility for the decision to submit for publication.

## CONTRIBUTORS

WL designed the study. TGS and LKT performed the cell manufacturing. YCL and MSS prepared the cell collection and clinical summary. YCL, JGL, JL, WJC, LPK, MLP, TTT and MSS participated in donor evaluation and eligibility screen. MC performed HLA analyses. KPN and KCH did the TCR typing. MSS coordinated regulatory affairs and activities across all sites. WL drafted the manuscript. YCL, TGS, LKT, JGL and MSS revised the manuscript. All authors participated in the interpretation of data, last revision of the manuscript, and approval of the final version.

## ACKNOWLEDGMENTS

This work was supported by a SingHealth Duke-NUS Academic Medicine COVID-19 Rapid Response Research Grant. We thank the donors for their blood donation, the nursing staff at Singapore General Hospital Haematology Center for sample collection, and Kee Chong Ng, Alex Tiong-Heng Sia, Yoke Hwee Chan, Germaine Liew, Shui Yen Soh, Kai-Qian Kam, Queenie Gan, Wilson Low and the staff at KK Research Centre for their administrative and technical supports.

---

**Supplemental Table 1.**
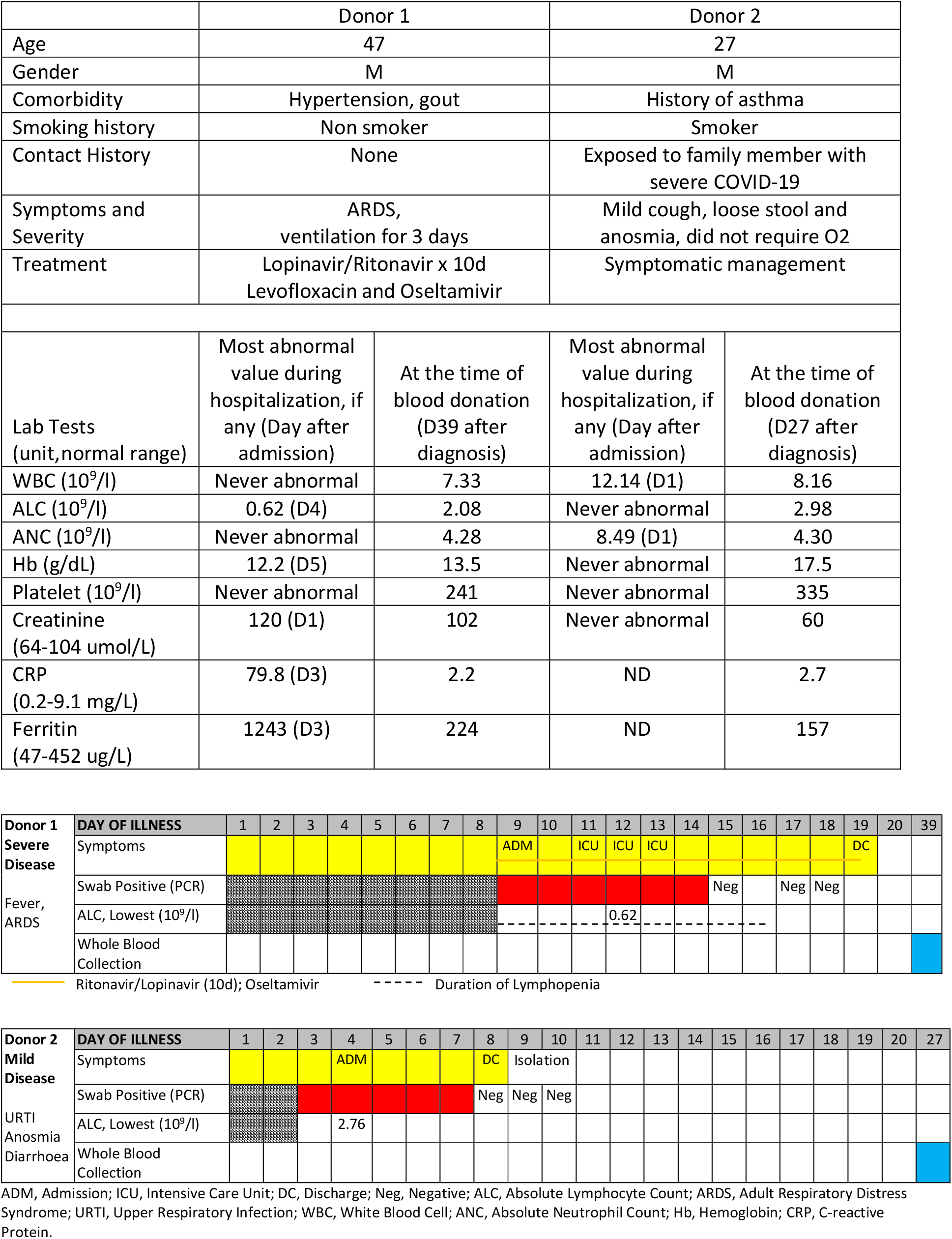
Medical Background

**Supplemental Table 2.** Phenotype Frequency in Singapore general population in relation to the two donors.

| HLA (Donor 1) | Chinese | Malay | Indian | HLA (Donor 2) | Chinese | Malay | Indian |
| --- | --- | --- | --- | --- | --- | --- | --- |
| A*26:01 | 4.0% | 1.2% | 8.4% | A*11:01 | 43.6% | 38.2% | 31.8% |
| A*33:03 | 18.8% | 13.7% | 25.5% | A*24:02 | 23.1% | 30.6% | 22.2% |
| B*08:01 | 2.2% | 0% | 6.1% | B*15:02 | 10.3% | 28.6% | 3.7% |
| B*58:01 | 20.1% | 9.4% | 6.1% | B*58:01 | 20.1% | 9.4% | 6.1% |
| C*03:02 | 25.7% | 7.7% | 6.1% | C*03:02 | 25.7% | 7.7% | 6.1% |
| C*07:02 | 39.6% | 23.3% | 15.4% | C*08:01 | 18.6% | 39.0% | 3.8% |
| DRB1*03:01 | 16.3% | 9.0% | NA | DRB1*03:01 | 16.3% | 9.0% | NA |
| DRB1*14:54 | NA | NA | NA | DRB1*04:05 | 13.1% | 1.8% | NA |
| ≥1 HLA-sharing<sup>#</sup> | 39.1% | 23.9% | 33.9% | ≥1 HLA-sharing<sup>#</sup> | 96.1% | 79.6% | 54.0% |
| Haplotype-sharing<br>(≥3 HLA)<sup>##</sup> | 13% | NA | NA | Haplotype-sharing (≥3 HLA)<sup>##</sup> | 21% | NA | NA |

Allele phenotype frequency = $1 - (1 - \text{allele frequency})^2$ ; Haplotype phenotype frequency = $1 - (1 - \text{haplotype frequency})^2$ , with frequencies obtained from www.allelefrequencies.net and ref 22,23.
<sup>#</sup>HLA-sharing is estimated conservatively summing the phenotype frequencies of HLA-A and -DR only, which have the least linkage disequilibrium, and excluding HLA-B and -C.
<sup>##</sup>A\*33:03 – C\*03:02 – B:58:01 – DRB1\*03:01; A\*11:01 – C\*08:01 – B:15:02; and C\*03:02 – B:58:01 – DRB1\*03:01 are common haplotypes among Chinese Singaporeans, with haplotype frequency at 7.0%, 3.4%, and 7.7%, respectively.

